# Etiological Factors of *Risk for Occupational Illness* in Nursing Professionals: Etiology and Risk Review Protocol

**DOI:** 10.1101/2024.09.29.24314573

**Authors:** Cyntia Leenara Bezerra da Silva, Ana Clara Dantas, Bárbara Ebilizarda Coutinho Borges, Leandro Melo de Carvalho, Heloise Rosália de Carvalho Nascimento, Maria Clara Regis de Souza, Allyne Fortes Vitor

## Abstract

**Aim:** To describe the protocol for a systematic review of etiology and risk to identify the etiological factors of *Risk for Occupational Illness* in nursing professionals.

**Design:** Etiology and risk review protocol, conducted according to JBI guidelines.

**Methods:** This is an etiology and risk review protocol to be carried out following the JBI guidelines in the following data sources: CINAHL, MEDLINE/PubMed, Cochrane Library, Scopus, EMBASE, Latin American and Caribbean Literature in Health Sciences, National Institute of Security and Health at Work and National Institute of Health at Work. The inclusion criteria are studies that address workers aged 18 years until retirement, demonstrate etiological factors that condition the susceptibility of the ND in question and address analyses regarding the identification, definition and association of factors with the *Risk of Occupational Illness*. The exclusion criteria are studies that did not address the topic, opinion articles, letters to the editor and editorials.

**Expected Results:** The results of this review will be publicly disclosed in scientific journals in the health area and will contribute to increasing the level of evidence of the Nursing Diagnosis *Risk for Occupational Illness*, so that they remain in future editions of NANDA International and that the diagnosis is updated and revised based on the current scientific literature.

**Conclusion:** It is expected identify the possible uses of the concept; determine the essential critical attributes; identify the antecedents and consequences of the concept; and define the empirical references and to highlight the etiological factors that are consistent with susceptibility to occupational illness.

**Strengths and limitations of this study:**

- This study will review the scientific literature on the etiological factors of *Risk for Occupational Illness* in nursing professionals, providing an increase in the level of evidence for this Diagnosis;
- The steps of this review will be followed strictly in accordance with the criteria of the JBI Manual for Evidence Synthesis;
- To avoid selection bias, data collection will occur independently with active blinding of researchers, who will use software to export and select data.

## Introduction

Workers’ health encompasses the interactions between work and the health-disease process, with activities regulated by the Organic Health Law, Law nº. 8080/90. This law shows that the State must provide favorable conditions for the full exercise of the work process. [1]

Therefore, economic and social policies are needed that propose the reduction of risks of diseases and other harm to professionals. In this context, the standard provides for the conditions for the execution of healthcare through health surveillance, the promotion, protection and recovery of workers’ health. [1]

Nursing professionals continuously provide health care 24 hours a day, sometimes due to a shortage of supplies and in precarious working conditions. Workers work 30-40 hours a week and, in some situations, have more than one employment relationship. [2] These facts, combined with the on-call regime, increase the tiring routine and interpersonal-professional conflicts that have repercussions on the biological and psychological dimensions, corroborating occupational illnesses that are largely neglected by the professionals themselves.

In general, healthcare professionals naturally present risk factors that are intensified depending on the conditions they are offered during the execution of care. For example, many healthcare institutions are considered unhealthy because they admit patients with a wide range of illnesses and require procedures that pose risks of accidents and illnesses to the worker. [2]

In Brazil, from 2018 to 2022, 329,176 work accidents related to exposure to biological material were recorded, of which 54.4% were nursing professionals. [3] Another important event to mention is the increasing number of accident benefits granted to workers, as a result of serious injuries, musculoskeletal diseases and mental disorders. [4]

Data from the International Labor Organization (2021) indicate that monetary expenses related to work-related accidents can account for up to 4% of the world’s Gross Domestic Product (GDP). This has the potential to be minimized or even eradicated, as many of these events are preventable as long as efficient risk management methods are implemented. [4]

Professional practice activities can potentially cause injury to workers and their occurrence is influenced and intensified by probability and severity. Awareness that occupational exposure to risks occurs in the face of professional-work environment interaction will allow nurses to develop and adopt a preventive mindset conducive to implementing strategies to minimize the possible risks of Occupational Illness. [2,4]

In order to develop a care plan that is appropriate for implementing nursing interventions and achieving expected results, nurses must rely on the Nursing Process (NP). The NP allows the identification, understanding, measurement and prediction of the individual’s health problems so that an effective and applicable care plan can be developed. Organized in five interrelated, interdependent, recurring and cyclical stages: assessment, diagnosis, outcome/planning, implementation, and evalution. [5-6]

In the NANDA-International (NANDA-I) taxonomy, the Nursing Diagnosis (ND) *Risk for Occupational Illness* (00404) approved in 2023, belonging to Domain 11. Safety/protection and Class 4. Environmental risks, presents 29 risk factors, classified as individual: (1) Difficulty with decision-making, (2) Excessive stress, (3) Improper use of personal protective equipment, (4) Inaccurate follow-through of employee health protocol, (5) Inaccurate follow-through to safety protocol, (6) Inadequate action to address modifiable factors, (7) Inadequate communication skills, (8) Inadequate knowledge of modifiable factors, (9) Inadequate social support, (10) Inadequate understanding of the importance of personal protective equipment, (11) Inadequate vaccination, (12) Inattentive to ergonomic principles, (13) Ineffective weight management. [7]

As well as, environmental factors: (1) Conflicted labor relationships, (2) Excessive workload, (3) Exposure to chemical agents, (4) Exposure to biological agents, (5) Exposure to intermittent impacts, (6) Exposure to psychosocial agents, (7) Exposure to repetitive motion activities, (8) Inadequate access to personal protective equipment, (9) Inadequate adoption of the ergonomic principle, (10) Inadequate biological monitoring, (11) Inadequate dosimetry monitoring, (12) Inadequate employee health protocol, (13) Inadequate placement of collective protective equipment, (14) Inadequate safety protocol, (15) Ineffective workload management, (16) Pathogen exposure. [7]

Highlighting at risk population: (1) Chestfeeding individuals, (2) Individuals whose work is comprised of monotonous activities, (3) Individuals with history of physical trauma, (4) Individuals with history of traumatic professional exposure, (5) Individuals with history of work-related accidents, (6) Individuals with limited access to healthcare services, (7) Individuals with multiple employment contracts, Individuals with responsibilities beyond own work ability, (9) Individuals with work-life imbalance, (10) Pregnant individuals, (11) Rotating shift workers. [7]

This ND has the definition: susceptibility to work-related condition or disorder resulting from a non-instantaneous event or exposure, and presents level of evidence 2.1, the initial level of a diagnosis approved to compose the taxonomy. [7]

Although NANDA-I presents itself as an estimated classification system, proposing through diagnostic classifications, the standardization of language among nursing professionals, it requires that constant theoretical, clinical and content studies be carried out to update the evidence and validity of diagnoses, to allow the expansion of the use of taxonomy in the most diverse scenarios and populations. [8]

The Nursing Diagnosis validation process is divided into three phases: concept analysis, content analysis by experts and clinical validation. The Etiology and Risk review will be necessary for the development of the concept analysis, the first stage of validation. [9]

## Aims

From this perspective, the following research question was raised: What are the etiological factors of the *Risk for Occupational Illness* in nursing professionals?

## Method

### Study

This is an Etiology and Risk Review protocol developed according to the recommendations of the JBI Manual for Evidence Synthesis, Chapter 7, and the PRISMA-ScR (Preferred Reporting Items for Systematic Reviews and Meta-Analyses extension for Scoping Reviews), registered in the International prospective register of systematic reviews with the respective registration: CRD4202549181. [10-11] The following steps will be adopted: definition of the research question, identification of relevant studies, assessment of methodological quality, data extraction and synthesis of results. [12]

This systematic literature review will be carried out with the aim of operationalizing the development of a concept analysis for the Nursing Diagnosis *Risk for Occupational Illness*. The review will be based on the acronym PEO, which are: P (population); E (etiology); and O (outcome). [10] Thus, the elements to be admitted will be P (nursing professionals); E (etiological factors); and O (*Risk for Occupational Illness*). To be adopted as the research question: “What are the etiological factors related to the Nursing Diagnosis *Risk for Occupational Illness* in nursing professionals?”

### Information sources

Searches in the literary collection will be carried out through the CAPES Periodicals Portal, through the Federated Academic Community (CAFe) platform of the Federal University of Rio Grande do Norte (UFRN) in the respective data sources: CINAHL (EBSCO), MEDLINE/PubMed (via National Library of Medicine), Cochrane Library, Scopus (Elsevier), EMBASE and Latin American and Caribbean Literature in Health Sciences (LILACS). As well as in the National Institute of Security and Health at Work (INSST) and National Institute of Health at Work (INST) for comparison between the Brazilian and Spanish scenarios, since Spain has intense discussion and visibility on the subject. It is worth noting that no restrictions on language or year of publication will be applied.

### Search strategy

The selection of descriptors used in the search strategy was done by consulting the Medical Subject Headings (MeSH) and the Health Sciences Descriptors (DeCS). Seven descriptors and one keyword related to the phenomenon were defined, in order to contribute to the findings of the available literature, according to Table 1.

**Table 1.** Description and characterization of the PEO mnemonic used in the research.

| **MNEMONIC**<br>**STRATEGY**<br>**PEO** | **DESCRIPTORS**<br>**DeCS** | **DESCRIPTORS**<br>**MESH** | **KEYWORD** |
| --- | --- | --- | --- |
| **P:** Nursing Professionals | "Nursing Professionals" | "Nursing professionals"<br>"Nursing Team" | - |
| **E:** Etiological Factors | "Etiology" | "Causality" "Etiology" | - |
| **O:** Risk for occupational illness | "Nursing diagnosis"<br>"Occupational risks"<br>"Occupational exposure"<br>"Occupational stress"<br>"Occupational diseases" | "Nursing diagnosis"<br>"Occupational risks"<br>"Occupational exposure"<br>"Occupational stress"<br>"Occupational diseases" | "Risk for occupational illness" |

The Boolean operators “AND” and “OR” were applied for the purpose of crossing the elements referenced by the acronym PEO. [13] The result of crossing the components of the acronym PEO with the Boolean operators was: “Nursing professionals” OR “Nursing Team” AND “Causality” OR “Etiology” AND “Nursing diagnosis” OR “Occupational risks” OR “Occupational Exposure” OR “Occupational stress” OR “Occupational diseases” OR “Risk for occupational illness”.

Considering the particularity of each data source, aiming at the greatest location of scientific evidence, cross-referencing of descriptors was strategically generated, resulting in product syntaxes (Appendix A).

### Inclusion criteria

As eligibility criteria, studies that address workers from 18 years of age until retirement, demonstrate etiological factors consistent with the susceptibility of the ND in question, and address analyses regarding the identification, definition and association of factors with the *Risk of Occupational Illness* will be considered. Studies that exclusively present etiological factors related to nursing professionals and that do not answer the research question will be excluded. Duplicate studies will be counted only once.

### Selection of sources of evidence

In order to ensure methodological quality and avoid selection bias, the selection of studies will be carried out independently and under active blinding of the two researchers using the Rayyan – Intelligent Systematic Review software (https://rayyan.ai/).

During the search process, all studies will be evaluated and their inclusion in the sample will occur after an initial reading of the title and abstract. In case of disagreement, a third reviewer will be consulted. The entire process follow the PRISMA-ScR flowchart, and will be properly documented detaileding of the selection process.

The methodological content of the studies found will be assessed by applying the checklists for studies available in the JBI manual for each type of study to be found in the review. [14] In the absence of a cut-off score established by the JBI, the decision on whether to include or remove a study from the sample will follow the cut-off score established by the researchers as ≥6. It should be noted that the score for each type of study will be measured based on the arithmetic mean of the number of items in each list.

### Data extraction

Data mapping will be performed using a structured data extraction tool in Microsoft Excel 2019. It will present variables related to the characterization of the studies, essential attributes, associated conditions and at risk population for the Nursing Diagnosis Risk for occupational illness. In addition, the identification of which etiological factors and at risk populations evidenced in the studies are not present in NANDA-I (Appendix B).

### Summary of results

In the summary of results, the findings identified by repeated reading of the studies will be grouped into categories developed by the researchers according to similarity of concepts. They will be presented in a descriptive manner, with the possibility of summarizing in tables, charts and other graphic resources in addition to the discursive text.

### Analysis of evidence

The studies that will make up the sample will undergo an analysis of the level of evidence, following the guidelines of the JBI approach classification: levels of evidence, where the following will be evaluated: level 1 – experimental studies; level 2 – quasi-experimental studies; level 3 – analytical observational studies; level 4 – descriptive observational studies; level 5 – expert opinion and bench research. [14] This analysis will fairly discuss the results of the studies according to the level of evidence presented.

### Ethical Aspects

Considering the nature of the theoretical research and the non-incorporation of human beings, submission to the Research Ethics Committee will not be necessary, since the scientific evidence to be used is available in the public domain.

## Results

The results will include the characterization of the studies in the sample, highlighting the etiological factors of the ND Risk for Occupational Illness in nursing professionals.

To this end, the results will be presented descriptively using tables and charts, in order to answer the research question. The studies in the sample will be organized by figure according to the Preferred Reporting Items for Systematic Reviews and Meta-Analyses.

## Discussion

The motivation for developing this review is based on the need to contemplate steps 3, 4, 7 and 8 of the concept analysis for the diagnosis of *Risk of Occupational Illness*, which are named as: identification of possible uses of the concept; determination of essential critical attributes; identification of antecedents and consequences of the concept; and definition of empirical references. [15] To clarify a theoretical gradient with the potential to revise the Nursing Diagnosis in NANDA-I, understanding the risks likely to threaten the safety and health of the worker, to attribute visibility to an individual beyond the patient and companion, the nursing professional.

Over the years, nursing has prospered, increasingly expanding the possibilities of professional practice and increasingly establishing itself as one of the main health sciences. The growing number of professionals reinforces the need for standardization of language among members of the nursing team, a fact made possible through the use of taxonomies, for example, NANDA-I.

However, for taxonomies to corroborate evidence-based practice, new research on *Risk of Occupational Illness* is required, impacting advances in knowledge regarding the state of the art of the object under study associated with the validation of the ND in NANDA-I so that its level of evidence is high. [7]

The process of validating nursing diagnoses is divided into the following phases: concept analysis, content analysis by experts, and clinical validation. Initially, the essential attributes, antecedents, and consequences of a Nursing Diagnosis are identified. After the concept analysis, the content will be evaluated by experts, proving its relevance, coherence, clarity, and relevance, so that it can be validated in clinical scenarios in the future. [9]

The definition and knowledge about indicators may result in greater health care coverage in terms of identifying responses and interventions in a timely manner with coherent and effective activities. [7,9,16] It is also evident that the identification of risk factors will enable the establishment of interventions that are consistent and targeted at the vulnerability presented by the worker, since, given the probability of prevention, any and all circumstances that make the professional susceptible to occupational illness will be recognized.

The results of this review will be publicly published in scientific journals in the health field and will contribute to increasing the level of evidence for the Nursing Diagnosis *Risk of Occupational Illness*, so that they remain in future editions of NANDA International and that the diagnosis is updated and revised based on current scientific literature.

## Conclusion

By identifying the possible uses of the concept; determining essential critical attributes; identifying antecedents and consequences of the concept; and defining empirical references, it will be possible to highlight the etiological factors (associated conditions, at risk population, and risk factors) consistent with susceptibility to occupational illness.

Identifying risk factors will enable the establishment of interventions that are consistent and targeted to the vulnerability presented by the worker, since, given the probability of prevention, any and all circumstances that make the professional susceptible to occupational ilness will be recognized. Beneficially impacting the health of the worker and, consequently, optimizing care, reflecting in a more well-founded care process and in the safety of those involved.

## Data Availability

All relevant data are within the manuscript and its Supporting Information files.

https://www.crd.york.ac.uk/prospero/

## Conflict of interest

None

## Supporting information

S1 Table. Table 1. Description and characterization of the PEO mnemonic used in the research.

S1 File.Appendix for Search strategies for each database and extraction instrument.

## FUNDING/ACKNOWLEDGEMENT

Coordination for the Improvement of Higher Education Personnel - doctoral scholarship for Dantas AC, Borges BEC and Carvalho LM and master’s scholarship for Silva CLB; the National Council for Scientific Technological Development for Vitor AF under funding code 3061062022-1; and by the Universal CNPq project no. 408730/2023-4.

## Notes

### Competing Interest Statement

The authors have declared no competing interest.

### Funding Statement

Yes

